# Post-COVID-19 Perceived Stigma-Discrimination Scale: Psychometric Development and Evaluation

**DOI:** 10.1101/2021.06.06.21258253

**Authors:** Carlos Arturo Cassiani-Miranda, John Carlos Pedrozo-Pupo, Adalberto Campo-Arias

**Author notes:** Correspondence to Adalberto Campo-Arias, Programa de Medicina, Facultad Ciencias de la Salud, Universidad del Magdalena, Carrera 32 No 22-08, Santa Marta (470004), Colombia. Teléfono: 57 5 438100, extensión 1338. **Contributors**: Carlos Arturo Cassiani-Miranda contributed to the study conception, and data interpretation and statistical analysis, drafted the article, and revised and approved the final version. John Carlos Pedrozo-Pupo and Adalberto Campo-Arias contributed to the design and study conception, data interpretation, and revised the intellectual content and approved the final version. **Data availability statement**: The data that support the findings of this study are available from the corresponding author upon reasonable request.

## Abstract

The COVID-19 survivors face social stigmatization, even with negative tests. Valid and reliable instruments are required to quantify the stigma-discrimination complex associated with COVID-19 (COVID-19-CED). The study aimed to adapt and evaluate a scale to measure COVID-19-CED in COVID-19 survivors. A validation study was done with 330 COVID-19 survivors between 18 and 89 years. The COVID-19 Perceived Discrimination Scale (C-19-PDS) was used, which was adapted from the Tuberculosis Perceived Discrimination Scale. An exploratory factor analysis (EFA), confirmatory factor analysis (CFA), internal consistency, and differential item functioning (DIF) were performed using the classical theory of tests. EFA showed a one-dimensional solution for the items of C-19-PDS; however, CFA showed poor goodness-of-fit indicators. The 5-item version of the C-19-PDS showed better goodness-of-fit indicators, high internal consistency, and non-gender DIF. In conclusion, the 5-item version of the C-19-PDS is one-dimensional, with high internal consistency, and without gender DIF. This instrument is recommended to evaluate COVID-19-CED in the Colombian population.

---

Despite the high physical, psychological and social-emotional burden experienced by people with COVID-19, these people face other problems after recovery, generating tremendous suffering (Liyanage-Don et al., 2021; Maheshwari et al., 2021). Psychological distress is present, even in asymptomatic or had few and mild symptoms during infection, which affects the quality and enjoyment of life (Balachandar et al., 2020).

Recent studies report that COVID-19 survivors face social stigma, even after complete remission, with negative tests for the virus (Dar et al., 2020; Mahmoudi et al., 2021). However, the stigmatization of COVID-19 survivors has not been systematically evaluated (Li et al., 2020; Maheshwari et al., 2021).

The stigma-discrimination complex related to COVID-19 (COVID-19-SDC) is associated with depressive complaints (Yuan et al., 2021), psychotic symptoms (Baral et al., 2021), and high suicidal risk (Campo-Arias et al., 2021). A routine evaluation of COVID-19-SDC should be carried out among COVID-19 survivors; this strategy is needed to measure the frequency of the phenomenon and implement the necessary actions to mitigate the negative impact (Li et al., 2020; Yuan et al., 2021). Valid and reliable instruments to quantify COVID-19-SDC are necessary to implement among the growing population of COVID-19 survivors (Dar et al., 2020). To date, there are no scales to evaluate COVID-19-SDC in this population.

The objective of the present study was to carry out the adaptation and psychometric evaluation of a scale to measure COVID-19-SDC in a Colombian sample of COVID-19 survivors.

## Method

### Design and participants

A validation study was designed with the participation of 330 COVID-19 survivors. They were aged between 18 and 89 years (Mean = 47.67, SD = 15.17); 61.52% were women and had a university education. The sample size was adequate for exploratory and confirmatory factor analysis since it is recommended to have 20 participants for each item (MacCallum et al., 2001). Table 1 presents demographic and clinical information on the participants.

**Table 1.** Sample description (N = 330).

| Variable | Category | n | % |
| --- | --- | --- | --- |
| Age (years) | 18–59 | 263 | 79.70 |
|  | 60 or more | 67 | 20.30 |
| Gender | Female | 203 | 61.52 |
|  | Male | 127 | 38.48 |
| Education | Primary | 29 | 8.79 |
|  | Secondary | 95 | 28.79 |
|  | University | 206 | 62.42 |
| Family income | Low | 235 | 71.21 |
|  | High | 95 | 28.79 |
| Marital status | Married or civil partnered | 218 | 66.06 |
|  | Single and others | 112 | 33.94 |
| Healthcare worker | Yes | 47 | 14.24 |
|  | No | 283 | 85.76 |

### Instrument

The COVID-19-SDC was explored with the COVID-19 Perceived Discrimination Scale (C-19-PDS). Ten of the eleven items of the Perceived Tuberculosis-Related Discrimination Scale were revised and adapted. Each item offers as response options: never (0), sometimes (1), often (2), and always (3) (Van Rie et al., 2008). According to current recommendations, the translation and back-translation process were carried out (Ramada-Rodilla et al., 2013). See annex 1.

### Procedure

In a pulmonology outpatient clinic at three institutions in Santa Marta, Colombia, the surviving COVID-19 were invited to participate in the study. The inclusion of participants was completed between October 12, 2020, and April 30, 2021. 70% (n = 231) of the patients were attended by teleconsultation and 30% (n = 99) in person. All participants self-completed an online questionnaire that was sent to the cell phone.

### Data analysis

#### Dimensionality

Exploratory factor analysis (EFA) was performed to find factor loadings and identify the items with the best performance. These loads are interpreted as other correlations and indicate the relationship between the item and the factor (Hefet & Liberman, 2017). Satorra-Bentler chi-square, Root Mean Square Error of Approximation (RMSEA) with 90% confidence interval 90% (90%CI), Comparative Fit Index CFI), Tucker-Lewis index (TLI), and Standardized Mean Square Residual (SMSR) were calculated in the confirmatory factor analysis (CFI). Satorra-Bentler chi-square was expected to show a probability greater than 0.05 or a ratio X^2^ / df < 5 (Bentler, 1976; Carmines & McIver, 1981), RMSEA ± 0.05, SMSR ≥ 0.05, and CFI and TLI values ≤ 0.90. The theoretical model with three acceptable coefficients is accepted (Hu & Bentler, 1999). The CFA was carried out in the Factor Analysis program.

#### Internal consistency

Internal consistency was calculated with the coefficients of Cronbach’s alpha (1951) and McDonald’s omega (1970). McDonald’s omega is a better indicator of homogeneity when the items show significant differences in factor loadings (Campo-Arias & Oviedo, 2008). The internal consistency must be between 0.70 and 0.95 (Keszei et al., 2010). These coefficients were calculated in Jamovi version 1.2.27.0.

#### Differential item functioning (DIF)

The gender DIF was quantified with Kendall’s tau-b (Kendall, 1938). Gender DIF was considered those correlations ≤ 0.20 (Hambleton, 2006). These calculations were performed in the SPSS version 23 program.

### Ethical issues

The research ethics board of the Universidad del Magdalena, Santa Marta, Colombia, approved the study (Act 002 of March 26, 2020). A free-use instrument was applied. Participation was voluntary, no incentives were offered, and informed consent was signed under national and international standards for research (World Medical Association, 2018).

## Results

### EFA and CFA

The EFA showed that the ten items best represented a one-dimensional solution; however, CFA showed poor goodness-of-fit indicators. The exploration of versions with fewer numbers showed acceptable goodness-of-fit indicators for a version with four and another with five items. Table 2 shows that the best solution is the version with five items. The commonalities and loadings for the five-item version are presented in Table 3.

**Table 2.** Goodness-of-fit indicators for the versions of 4-, 5- and 10-items.

| Indicator | Four items<br>(1, 3, 4, and 5) | Five items<br>(1, 3, 4, 5, and 9) | Ten items |
| --- | --- | --- | --- |
| $\chi^2$ (df) | 14.22 (2) | 16.34 (5) | 465.90 (35) |
| $\chi^2 / df$ | 7.11 | 3.27 | 13.31 |
| CFI | 0.98 | 0.99 | 0.78 |
| TLI | 0.95 | 0.97 | 0.71 |
| RMSEA (90CI%) | 0.14 (0.08 – 0.21) | 0.08 (0.04 – 0.13) | 0.19 (0.18 – 0.21) |
| SRMR | 0.02 | 0.02 | 0.11 |
| Cronbach's alpha | 0.87 | 0.83 | 0.85 |
| McDonald's omega | 0.88 | 0.86 | 0.87 |

**Table 3.** Commonalities, loadings, and Kendall’s tau b.

| Item | Commonality | Loading | Kendall's tau b<sup>1</sup> |
| --- | --- | --- | --- |
| 1. I am still contaminated | 0.71 | 0.84 | 0.04 |
| 3. Some people avoid me | 0.82 | 0.91 | 0.02 |
| 4. People recommend their family avoid me | 0.67 | 0.82 | 0.07 |
| 5. Some people treat me badly | 0.39 | 0.62 | 0.01 |
| 9. I stay away from other people | 0.19 | 0.44 | 0.04 |

<sup>1</sup>Gender DIF.

### Internal consistency

The global scale and versions of four and five items showed acceptable internal consistency, with Cronbach’s alpha values between 0.83 and 0.87; McDonald’s omega of 0.86. See details in Table 2.

### Gender DIF

The five-item scale showed Kendall’s t between 0.01 and 0.07, indicating that the items are free of gender bias. See Table 3. In addition, it was observed that the scores were significantly higher in men than in women [2.84 (SD = 3.03) versus 2.61 (SD = 2.64), Levene’s test for equality of variances F = 0.02, p = 0.89, t = 0.69; df = 328, p = 0.49].

## Discussion

The versions of 10, 5, and 4 items of the C-19-PDS showed acceptable internal consistency. However, CFA showed that the 5-item version presented a one-dimensional structure with better goodness-of-fit indicators. This 5-item version of the C-19-PDS shows non-gender DIF.

All versions of the C-19-PDS showed an excellent internal consistency (Cronbach’s α between 0.83 and 0.87) comparable with the homogeneity of the original scale for perceived discrimination related to tuberculosis (Cronbach’s alpha between 0.82 and 0.91, McDonald’s omega was not reported) (Van Rie et al., 2008). McDonald’s omega is the best estimator of internal consistency when the tau-equivalence principle is violated (Campo-Arias & Oviedo, 2008). This similarity in the scales for the C-19-PDS and Perceived Tuberculosis-Related Discrimination Scale suggests high-reliability instruments (Keszei et al., 2010).

Poor goodness-of-fit indicators for the 10-item version of C-19-PDS invited testing of other versions. The 5-item version of the C-19-PDS preserves the one-dimensional, as expected, with better indicators than the Perceived Tuberculosis-Related Discrimination Scale (Van Rie et al., 2008). The shortened versions of the instruments have their advantages: they are usually one-dimensional and fit acceptably with the data. The factorial solution for the 5-item version of the C-19-PDS is encouraging (Campo-Arias & Oviedo, 2008).

The C-19-PDS had non-gender DIF. This observation suggests that higher total scores observed among men are independent of item bias. The total scores indicate fundamental differences in the response pattern in men and women (Hambleton, 2006).

### Practical implications

These findings should be considered preliminary. The 5-item version of the C-19-PDS had better dimensionality than the 4- and 10-item versions. However, the 4- and 10-item versions of the C-19-PDS may be helpful depending on the study’s objectives. These findings should be verified in new studies in Spanish and other languages. The Spanish version of the C-19-PDS can be used in clinical and epidemiological studies to evaluate COVID-19-SDC (Keszei et al., 2010).

This work contributes to a greater understanding and appropriate measurement of COVID-19-SDC (Campo-Arias, 2021b). A valid, reliable, and non-gender biased instrument helps measure the effect of interventions to reduce COVID-19-SDC in further researches (Cassiani-Miranda et al., 2020; Li et al., 2020; Yuan et al., 2021).

### Study strengths and limitations

This study presents a new instrument to evaluate COVID-19-SDC in Spanish speakers. However, this research has the limitation; it did not quantify the instrument’s stability (test-retest assessment), information necessary when repeated evaluations are made (Afhami et al., 2017). Likewise, it would be interesting to evaluate the scale performance with models based on the item response theory (Liu et al., 2019).

## Conclusions

The 5-item version of the C-19-PDS is a one-dimensional instrument with high internal consistency and without gender DIF. This instrument is recommended to evaluate COVID-19-SDC in the Colombian population. It is necessary to corroborate these findings in other Spanish-speaking countries and other languages and test the performance with models based on item response theory.

## Data Availability

The data that support the findings of this study are available from the corresponding author upon reasonable request.

## Acknowledgment of conflict of interest

The authors have no conflicts of interest to declare.

## Annex 1. The items of C-19-PDS

1. Some people think I am still contaminated because I had COVID^1^
2. Some people think that I can still infect them because I had COVID.
3. Some people avoid me since I had COVID.^1^
4. Some people recommend that their family members avoid contact with me because I had COVID.^1^
5. Some people treat me badly when they know I had COVID.^1^
6. I keep a secret that I had COVID.
7. I forbid my family members to comment that I had COVID.
8. I worry that my relatives are poorly treated because I had COVID.
9. I stay away from other people because I could still transmit COVID to them.^1,2^
10. I feel discriminated against because I had COVID.

^1^Items included in 5-item version.

^2^Item removed of 4-item version.

Scoring: (0) never, (1) sometimes, (2) often, and (3) always.

